# The Bacterial Profile and Antibiotic Susceptibility Pattern in Respiratory Tract Samples from ART-Experienced HIV-Positive Adults in Uganda

**DOI:** 10.1101/2023.02.28.23286566

**Authors:** Lubega Gloria, Abaasa Andrew, Willyfred Ochola, Bernard Kikaire, Joseph Lutaakome, Eugene Rugazira, Yunia Mayanja

## Abstract

**Introduction:** Microbial infections are a major cause of morbidity and mortality among people living with HIV (PLWH). Respiratory tract infections (RTIs) are responsible for approximately 70% of illnesses among PLWH. Drug resistant bacteria are highly prevalent among PLWH and this is a public health concern.

**Methods:** This is a retrospective analysis of data collected during the COSTOP trial between 2011 and 2013. Sputum collected on spot from participants presenting with a productive cough was examined using Gram, Ziehl-Neelsen stains and cultured on suitable bacteriological media. Antimicrobial sensitivity testing was done on isolated pathogens, by disc diffusion technique.

**Results:** We included 687 participants with mean age 41.3 (SD 8.2) years of whom 76.4% were female. Two hundred one sputum samples grew bacteria; *Moraxella species* (27.4%), *Streptococcus pneumoniae*(25.4%), *Haemophilus influenza*(22.4%), *Mycobacterium species*(4.5%), *Pseudomonas species*(4.0%), *Staphylococcus aureus*(4.0%), *Escherichia coli* (1.0%), *Klebsiella species* (1.0%), other bacteria (10.4%). A higher monthly income greater than or equal to 30$ (aOR= 0.63, 95%CI: 0.40-0.99) and longer duration since HIV diagnosis (aOR= 1.06, 95%CI: 1.0-1.11) were found to be independently associated with a positive bacterial culture. Moraxella *sp*, H. *influenza* and Pseudomonas had zero sensitivity towards cotrimoxazole. Sensitivity to erythromycin was low among Moraxella *sp* (28.6%), H. *influenza* (31.6%) and S. *aureus*(42.9%) and other bacteria (42.9%). Most isolates were sensitive to Amoxicillin + Clavulanic acid and ceftriaxone.

**Conclusion:** There is a very low sensitivity of isolated bacteria to commonly prescribed antibiotics that are more available through the national supply chain, which is of public health concern. Urgent steps to tackle the high antimicrobial resistance among PLWH is required.

## Introduction

For over three decades, Human Immunodeficiency Virus (HIV) infection remains a disease of public health importance with approximately 37.6 million people living with HIV (PLWH) globally in 2020 (1). Sub-Saharan Africa (SSA) suffers the highest burden of HIV with nearly 70% of global HIV infections (2). Since the introduction of antiretroviral therapy (ART), PLWH have an improved quality of life however, microbial infections are still a major cause of morbidity and mortality among this population (3), with approximately 70% of illnesses being respiratory tract infections (RTIs) (4, 5). Ojha et al. reported a high prevalence (47%) of respiratory tract infections caused by bacterial pathogens among PLWH(5) and, factors associated with these infections include low CD4 counts (<200 cells/µl) and detectable viral loads (5, 6).

HIV infection causes a progressive depletion of CD4 T cells as well as an impairment of cellular and humoral immunity through a dysfunction of the T and B cells respectively (7). A dysfunction of T cells leads to abnormal cellular responses while a dysfunction of B cells leads to a lack of antibody responses to infections. The resultant immune dysfunction, deregulation and depletion of CD4 lymphocytes causes an increased susceptibility to infections and the subsequent risk of other complications like resistant pathogens (8, 9). HIV infection causes an alteration in lung host defences for example, it affects mucociliary function which may contribute to an increase RTIs among PLWH (9). PLWH are at increased risk of hospital acquired infections due to their frequent contact with health care system through frequent clinic visits and admissions (10). The frequent infections and admissions among PLWH, pill burden leading to unfinished doses and inappropriate use of drugs through self-medication are some of the factors that have led to the development of antimicrobial resistance among this population (11).

Antimicrobial resistance is an emerging global problem of public health importance (4). Previous studies have reported a much higher prevalence of drug resistant bacteria among PLWH (79%) compared to their HIV negative counterparts (30%) (10, 12). Prophylactic cotrimoxazole given to PLWH over prolonged periods of time to prevent opportunistic infections has been reported to give rise to antibiotic resistance (13). Resistant microbes are more difficult to manage since they require alternative medications and or higher doses of drugs, both of which are more expensive, not readily available in SSA and or toxic to the patient (13). Additionally, laboratory capacity for identifying AMR is still limited in many parts of SSA; data are therefore still limited on antimicrobial susceptibility patterns among PLWH in SSA (10). (10). It is important to understand the common disease-causing pathogens among PLWH and current antibiotic susceptibility patterns for better management. We studied the susceptibility pattern of microorganisms isolated from respiratory samples taken from PLWH in central Uganda.

## Methods

### Study design, setting

We conducted a retrospective analysis of data collected during the COSTOP trial *(Safety of discontinuing cotrimoxazole prophylaxis among HIV infected adults in Uganda: a randomised controlled trial, ISRCTN44723643)* conducted between January 2011 and March 2013 in two HIV care clinics in Masaka and Entebbe in central Uganda. (14, 15).

The COSTOP trial enrolled HIV positive adults (≥18 years) who had been on ART and cotrimoxazole prophylaxis for at least six months, clinically asymptomatic and had CD4 cell counts of >250 cells/mm^3^ prior to enrolment. Individuals who had an acute illness, grade three or four anaemia, neutropenia or thrombocytopenia, known hypersensitivity to cotrimoxazole, and pregnant women were excluded.

### Participants and Eligibility

The current analysis included COSTOP trial participants who presented to the clinic with a productive cough and provided sputum at any time during the trial. Participants were asked to collect sputum in a dry, leak proof, sterile container after rinsing their mouth with water. Samples were immediately transported to the laboratory for microscopy, culture, and sensitivity.

### Laboratory

Samples were examined microscopically using Gram, Ziehl-Neelsen stains and cultured on suitable bacteriological media (Blood, Chocolate, MacConkey agar and Subouroid agar). Samples were incubated at 37^0^c for 24hrs, the pathogens that grew from incubated samples were identified microscopically and biochemically using analytical profile index (API) and by sensitivity testing. Antimicrobial sensitivity testing was done on isolated pathogens, by disc diffusion technique(16). The diameter of the zone of inhibition was measured and compared to that of European Committee on Antimicrobial Susceptibility Testing (EUCAST) standards(17) to determine resistance or sensitivity of a pathogen to the antimicrobial.

### Study variables

The dependent variable was bacterial culture positivity of sputum collected during the study. The independent variables were gender, age (years) categorized under age groups <35years, 35-55 years and >55 years, recent and initial CD4 counts categorized into <200, 200-300 and >300 cells/ul, study arm in the COSTOP trial (cotrimoxazole arm, placebo arm), highest education level attained categorized into none, primary and secondary school, marital status categorized into married, divorced and single, monthly income ($) categorized into those earning <30 and >/=30, ART regimen categorized into First line and second line, duration on ART (years) and period since HIV diagnosis (years).

### Statistical analysis

The data collected were managed in MS Access, 2003 (Microsoft Corporation, Redmond, WA) and data analyses were performed using Stata 14 (Stata Corp, College Station, Texas 77845 USA) [11]. Continuous variables were presented as means with standard deviation (SD) and categorical variables as frequencies and percentages. The proportion of bacterial culture positivity overall, and by different participant characteristics were determined as number culture positive divided by the total sample, expressed as a percentage. A bar graph was used to display the frequency of bacterial isolates, and frequency and percentages used to summarise antibacterial susceptibility of bacteria isolated in sputum. To determine factors associated with bacterial culture positivity, we fitted both univariable and multivariable logistic regression models. At univariable analysis, all factors in logit models with likelihood ratio test p-value <0.2, were selected for multivariable logit model. Factors were retained in the multivariable logit model if their inclusion did not make the model fit significantly worse at p<0.05 on likelihood ratio test. We present the results as adjusted odds ratios (AOR) with corresponding 95% confidence intervals (CI)

## Results

### Participants’ characteristics

We included 687 participants in the analysis with mean age was 41.3 years (SD ±8.23) and 76.4% were female. Overall, 51.1% of participants received cotrimoxazole, 70.2% earned less than $30 monthly and 60.3% had primary level education. We observed that the mean period since HIV diagnosis was 5.49 years (SD ±4.50) and the mean period on ART was 3.79 years (SD ±1.86) with 97.9% of participants on a first line ART regimen. Regarding CD4 counts, 89.7% had a recent CD4 of >300cell/µl however, 68.4% had a nadir CD4 count of <200cell/µl (Table 1).

**Table 1:** Factors associated with bacterial growth in sputum cultures among ART-experienced HIV-positive individuals in Uganda (N=687)

| Characteristic | N =687<br>n(%) | Culture positive<br>(%) | OR (95% CI) | P-value | AOR (95% CI) | P-value |
| --- | --- | --- | --- | --- | --- | --- |
| Sex |  |  |  |  |  |  |
| Female | 525 (76.4) | 113 (21.5) | 1 |  | 1 |  |
| Male | 162 (23.6) | 43 (26.5) | 1.32 (0.88-1.98) | 0.183 | 1.37 (0.89-2.11) | 0.156 |
| Age (years) |  |  |  |  |  |  |
| <35 | 161 (23.4) | 40 (24.8) | 1 |  |  |  |
| 35-55 | 482 (70.2) | 110 (22.8) | 0.89 (0.59-1.36) | 0.599 | 0.91 (0.59-1.41) | 0.666 |
| >55 | 44 (6.4) | 6 (13.6) | 0.48 (0.19-1.21) | 0.120 | 0.43 (0.17-1.13) | 0.086 |
| Receiving cotrimoxazole |  |  |  |  |  |  |
| cotrimoxazole | 351 (51.1) | 71 (20.2) | 1 |  |  |  |
| placebo | 336 (48.9) | 85 (25.3) | 1.34 (0.93-1.91) | 0.113 | 1.37 (0.94-1.98) | 0.098 |
| Education level |  |  |  |  |  |  |
| None | 101 (14.7) | 26 (25.7) | 1 |  |  |  |
| Primary | 414 (60.3) | 91 (22.0) | 0.81 (0.49-1.34) | 0.419 |  |  |
| Secondary | 172 (25.0) | 39 (22.7) | 0.85 (0.48-1.50) | 0.566 |  |  |
| Marital status |  |  |  |  |  |  |
| Married | 284 (41.3) | 62 (21.8) | 1 |  |  |  |
| Divorced | 377 (54.9) | 88 (23.3) | 1.09 (0.75-1.58) | 0.646 |  |  |
| Single | 26 (3.8) | 06 (23.1) | 1.07 (0.41-2.79) | 0.883 |  |  |
| Monthly income (\$) | | | | | | |
| <30 | 448 (70.2) | 109 (24.3) | 1 |  |  |  |
| >/=30 | 190 (29.8) | 38 (20.0) | 0.78 (0.51-1.18) | 0.236 | 0.63 (0.40-0.99) | 0.049 |
| Recent CD4* |  |  |  |  |  |  |
| <200 | 11 (1.6) | 1 (9.1) | 1 |  |  |  |
| 200-300 | 58 (8.4) | 18 (31.0) | 4.50 (0.53-37.85) | 0.166 | 5.09 (0.58-44.60) | 0.141 |
| >300 | 616 (89.7) | 137 (22.2) | 2.86 (0.36-22.54) | 0.318 | 2.86 (0.35-23.44) | 0.326 |
| Nadir CD4* |  |  |  |  |  |  |
| <200 | 470 (68.4) | 105 (22.3) | 1 |  |  |  |
| 200-300 | 123 (17.9) | 30 (24.4) | 1.12 (0.70-1.79) | 0.629 |  |  |
| >300 | 18 (2.6) | 4 (22.2) | 0.99 (0.32-3.08) | 0.991 |  |  |
| ART regimen* |  |  |  |  |  |  |
| First line | 662 (97.9) | 151 (22.8) | 1 |  |  |  |
| Second line | 14 (2.1) | 1 (7.1) | 0.26 (0.03-201) | 0.196 | 0.24 (0.03-1.94) | 0.180 |
| Duration on ART, (mean (SD)) yrs | 3.79 (1.86) | 3.75 (1.84) | 0.98 (0.89-1.08) | 0.726 |  |  |
| Years since HIV diagnosis, (mean (SD)) yrs | 5.49 (4.50) | 6.20 (6.83) | 1.04 (1.00-1.09) | 0.053 | 1.06 (1.01-1.11) | 0.025 |

ART = antiretroviral therapy; HIV = Human Immunodeficiency Virus

### Bacterial culture positivity and prevalence of isolated microorganisms

A total of 201 (29.3%) samples had bacterial growth, with 9 samples growing more than one bacterial organism. Isolated bacteria included *Moraxella species* (55, 27.4%), *Streptococcus pneumoniae* (51, 25.4%), *Haemophilus influenza* (45, 22.4%), *Mycobacterium species* (11, 4.5%), *Pseudomonas species* (8, 4.0%), *Staphylococcus aureus* (8, 4.0%), *Escherichia coli* (2, 1.0%), *Klebsiella species* (2, 1.0%) and other bacteria (19, 10.4%) (Figure 1).

**Fig.1.**
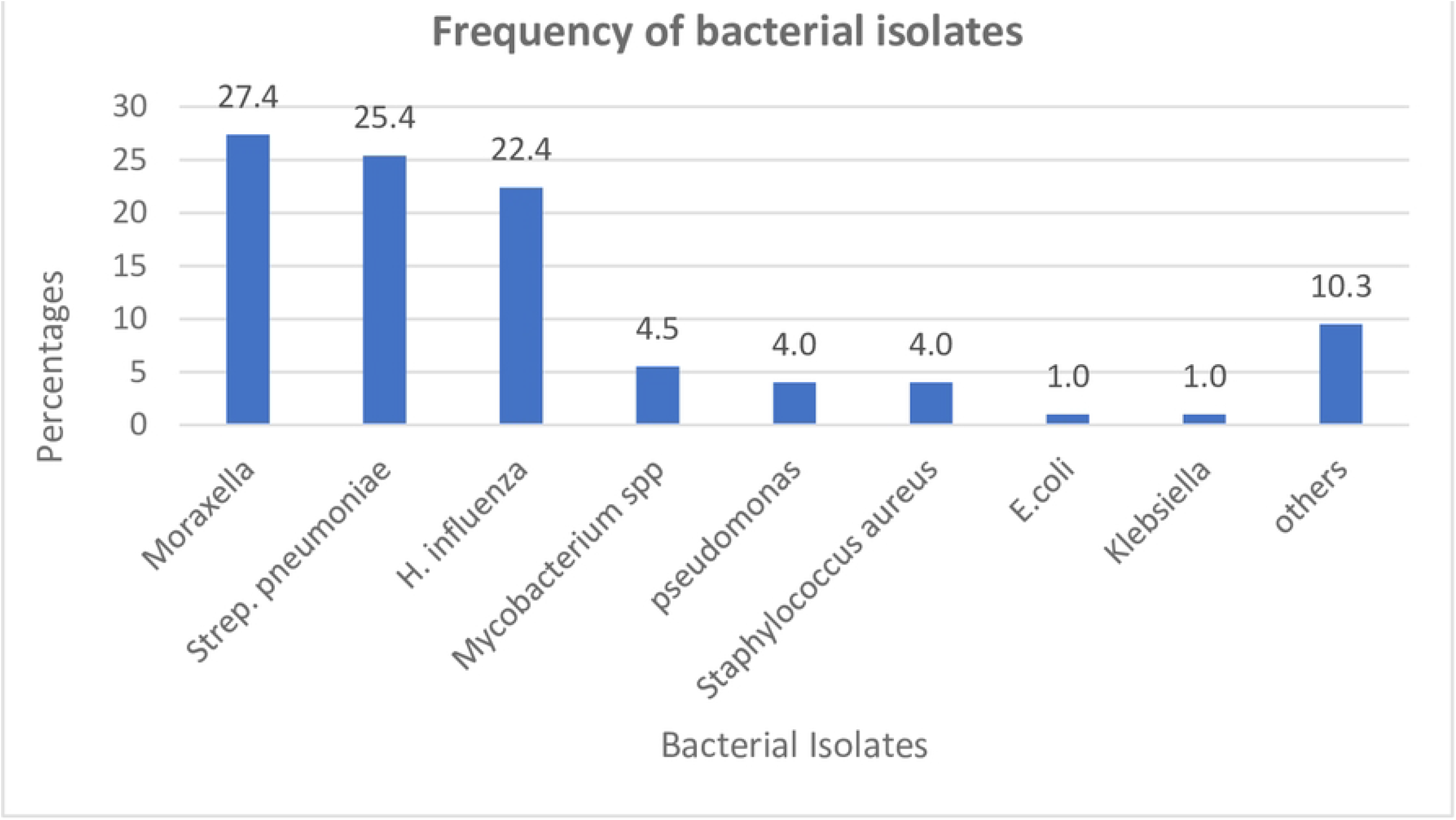
Frequency of bacterial isolates.

### Factors associated with bacterial culture positivity

The factors found to be independently associated with having a bacterial culture positive sample where; monthly income > $30 (aOR= 0.63, 95%CI 0.40-0.99) compared to those earning below $30 and longer duration of years since HIV diagnosis [aOR= 1.06, 95% CI 1.01-1.11]. Participants who received placebo were more likely to have a positive sputum culture than those who received cotrimoxazole however this was not found to have statistical significance (aOR= 1.37, 95% CI 0.94-1.98) (Table 1).

### Antibiotic sensitivity of isolated microorganisms

The sensitivity of bacterial isolates is shown in Table 2. Most isolates were highly sensitive to Amoxicillin + Clavulanic acid and ceftriaxone with *Moraxella sp. and, Streptococcus pneumoniae* having 100% sensitivity to both. *Pseudomonas* had 100% sensitivity to gentamycin while H. *influenza* showed 100% sensitivity to ceftriaxone and 88.6% sensitivity to Amoxicillin + Clavulanic acid. Other bacteria showed the highest sensitivity to ceftriaxone and gentamycin (85.7%).

**Table 2:**
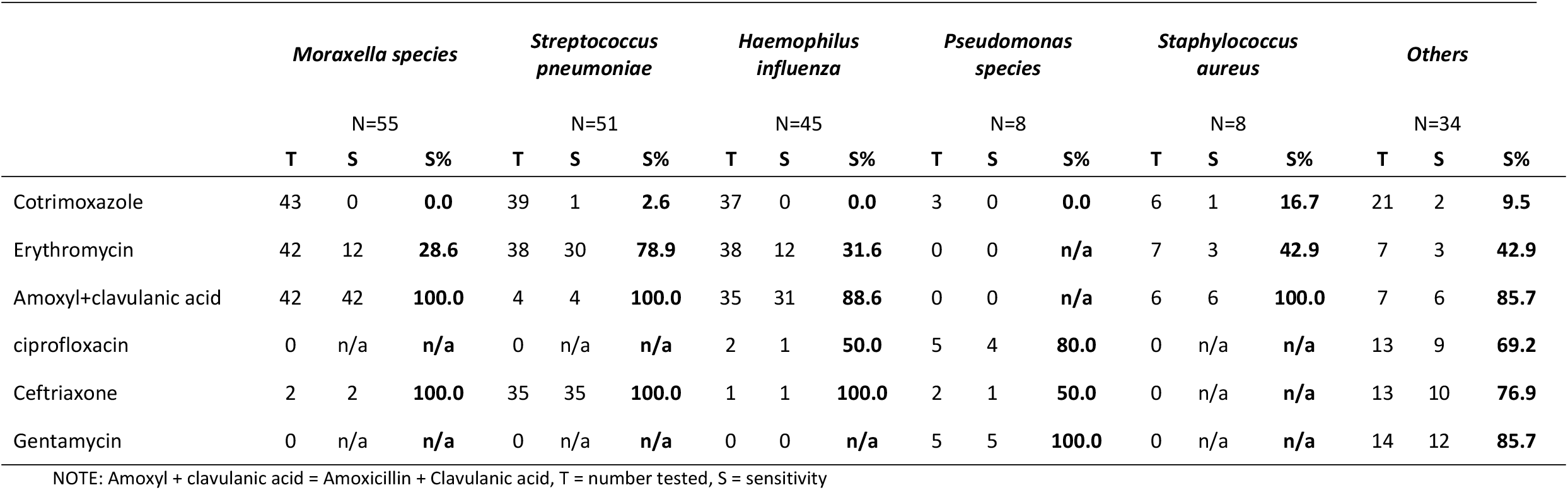
Antibacterial susceptibility rates of bacterial isolated from sputum of PLWH.

All isolates had very low sensitivity to cotrimoxazole (<16.7%). *Moraxella sp*, H. *influenza* and *Pseudomonas* showed zero sensitivity to cotrimoxazole. *Moraxella s*p (28.8%), H. *influenza* (31.6%), S. *aureus* (42.9%) and other bacteria (42.9%) also had low sensitivity towards erythromycin. H. *influenza* (50.0%) and other bacteria (69.2%) had relatively low sensitivity towards ciprofloxacin.

## Discussion

We cultured sputum from ART experienced PLWH with a productive cough and approximately 30% of the sputum samples had bacterial growth. This is similar to what was reported by Akingbade OA et al who found a prevalence of 24.2% positive cultures from sputum collected from participants with lower RTIs (18), but lower than approximately 44% prevalence reported by Adhanom et al (19). Adhanom et al included both ART experienced and ART naïve participants that were suspected of pneumonia, which more a severe disease than the presentation of cough that was considered in our study. Our participants were all ART experienced, indicating they might have had a better immunity than those included in Adhanom et al study. Additionally, Adhanoum et al’s study was conducted in three years after the COSTOP study, during which period there might have been an increase in antimicrobial resistance, leading to reduced response by the bacteria to antibiotics hence increased bacterial growth on culture.

We found a high prevalence of *Moraxella species* (28%) in our sputum samples. *Moraxella species* is usually normal flora of the respiratory tract, however, previous studies have reported *Moraxella* as a pathogen of the lower respiratory tract in immunosuppressed patients (20). Abdullah et al reported a relatively high prevalence of *Moraxella* in patients with lower RTIs among the elderly and immunosuppressed (20). Our finding of high proportion of participants with *Moraxella species* may indicate a less than complete immune recovery in these patients. The Moraxella species isolated in our study was sensitive to amoxicillin+ clavulanic acid and ceftriaxone and had very low sensitivity to erythromycin and cotrimoxazole, which is similar to what Abdullah et al reported (20). The very low sensitivity of all the bacterial isolates to cotrimoxazole is not surprising since our study participants were either on active cotrimoxazole prophylaxis, or had previously used cotrimoxazole prophylaxis.

HIV infection leads to a low CD4 count and a defective immune system which renders people infected with the HIV virus susceptible to microbial infections leading to a greater use of antibiotics among people living with HIV. The increased use of antibiotics may explain the observed low sensitivity of isolated bacteria to cotrimoxazole (21), and erythromycin (20) among PLWH in this study and other studies.

Participants that received placebo were more likely to have a positive sputum culture compared to those receiving cotrimoxazole, however this was not statistically significant. This implies that use of Cotrimoxazole did not provide protection against respiratory infections in this study population. This could be due to the fact that the study participants were ART experienced, had improved and stabilised immunity, and thus were no longer susceptible to infections associated with compromised immunity. Although the study was not powered to answer the question on the effect of cotrimoxazole on respiratory bacterial infections, this finding supports the 2020 Uganda HIV prevention and treatment guidelines that recommends use of prophylactic cotrimoxazole only in adults with advanced HIV disease, those newly diagnosed with HIV, children below 18years, pregnant and breastfeeding women(22). An association of long term use of cotrimoxazole among PLWH with an increase in resistance to cotrimoxazole has been reported in previous studies (23, 24) therefore, reduced use of cotrimoxazole in this population may reduce resistance of microbes to it.

The participants in the study were generally low earners as majority of them were earning less than $30 a month. Participants who earned more than $30 were less likely to have positive bacterial cultures compared to those who earned less than $30. This is similar to evidence that suggests that those with lower socio-economic status are more vulnerable, have higher burden of disease and thus more prone to frequent respiratory infections (25).

Participants who had a longer duration since HIV diagnosis were more likely to have a positive bacterial culture. This finding requires further investigation since there is no clear biological explanation between duration of HIV infection and risk of bacterial infections.

## Strengths and Limitations

This study among ART experienced participants has applicable results as majority of PLWH are on ART following the test and treat policy. Previous studies have mainly reported antimicrobial sensitivity among ART naïve patients. The study was however, a secondary data analysis and this limited the number of antimicrobial drugs that could have been tested for bacterial sensitivity. We could also not explore other factors for example viral load, that could be associated with a positive sputum culture.

## Conclusion

High prevalence of bacterial isolates with most being susceptible to Amoxicillin + Clavulanic acid and ceftriaxone. However, majority of organisms showed very low sensitivity to cotrimoxazole and erythromycin that are cheaper and readily available. As antibiotic choices are becoming limited due to low sensitivity of microbes to these drugs, there is an urgent need to invest in the prevention and control of increasing antibiotic resistance.

## Data Availability

All relevant data are within the manuscript and its Supporting Information files.

## Abbreviations

API: Analytical profile index
AMR: Antimicrobial Resistance
ART: Antiretroviral therapy
CD4: Cluster of Differentiation 4
COSTOP: Safety of discontinuing cotrimoxazole prophylaxis among HIV infected adults in Uganda: a randomised controlled trial
EUCAST: European Committee on Antimicrobial Susceptibility Testing
HIV: Human Immune Deficiency Virus
PLWH: People living with HIV
RTIs: Respiratory tract infections
SSA: Sub-Saharan Africa

## Acknowledgements

We would like to acknowledge the contributions of the COSTOP trial team and the contribution of the study participants

## Ethical approval statement

All participants enrolled into the main COSTOP trial gave written informed consent prior to any study procedures. The COSTOP study received approval from Uganda Virus Research Institute Ethics committee, National Drug Authority and Uganda National Council for Science and Technology.

## Author Contributions

LG conceived and designed the study, participated in data collection, analysis and interpretation, AA contributed to data analysis. All authors read, revised the original manuscript and approved the final version of manuscript.

